# Occupational risk of COVID-19 in the 1^st^ vs 2^nd^ wave of infection

**DOI:** 10.1101/2020.10.29.20220426

**Authors:** Karin Magnusson, Karin Nygård, Fredrik Methi, Line Vold, Kjetil Telle

**Affiliations:** Norwegian Institute of Public Health, Cluster for Health Services Research, Oslo, Norway; Lund University, Faculty of Medicine, Department of Clinical Sciences Lund, Orthopaedics, Clinical Epidemiology Unit, Lund, Sweden; Norwegian Institute of Public Health, Division of Infection Control and Environmental Health, Oslo, Norway

**Author notes:** **Contact information, corresponding author**: Karin Magnusson, Cluster for Health Services Research, Norwegian Institute of Public Health, Postboks 222, Skøyen, N-0213 Oslo, Visiting address: Sandakerveien 24c, Building D, 0473 Oslo.

## Abstract

**Aim:** To study whether employees in occupations that typically imply close contact with other people are tested more and at higher risk of confirmed SARS-CoV-2 infection (COVID-19) and related hospitalization, in the 1^st^ and 2^nd^ wave of infection in Norway.

**Methods:** In all 3 559 694 residents of Norway on January 1^st^ 2020 aged 20-70 (with mean [SD] age 44.1 [14.3] years and 51% men), we studied COVID-19 testing patterns sorted by occupation (using Standard Classification of Occupations [ISCO-08]). We also studied whether selected occupations had a higher risk of 1) confirmed COVID-19 and 2) hospitalization with COVID-19, compared to everyone else aged 20-70 years using logistic regression adjusted for age, sex, testing behavior, and own and maternal country of birth.

**Results:** Occupations with high frequency of testing (e.g. health personnel and teachers) had a low frequency of positive tests. Nurses, physicians, dentists, physiotherapists, bus/tram and taxi drivers had 1.1-4 times the odds of COVID-19 during the 1^st^ wave, whereas bartenders, waiters, transport conductors and travel stewards had 1.1-3 times the odds of COVID-19 during the 2^nd^ wave (when compared to everyone else). Teachers had moderately increased odds of COVID-19. Occupation may be of limited relevance for hospitalization with the disease.

**Conclusion:** Studying the entire Norwegian population using international standardized codes of occupations, our findings may be of relevance to national and regional authorities in handling the pandemic. Also, our findings provide a knowledge foundation for the more targeted future studies of lockdown, testing strategies and disease control measures.

## Introduction

The coronavirus disease 2019 (COVID-19) emerged in late 2019 in China and has in December 2020 resulted in over 70.000.000 infected and over 1.600.000 deaths globally [1]. In the Nordic countries, the first cases with confirmed SARS-CoV-2 infections probably originated from bars and restaurants in Austria and Italy when Nordic residents visited the countries during winter holidays February 2020 [2]. Later, testing, contact tracing and quarantine in addition to lockdown restrictions of activities particularly in trade, catering, travel and tourism industries are believed to have reduced the spread of the virus, whereas the lockdown of schools and pre-schools are assumed to have had a smaller effect [2-4]. However, to what extent occupational settings implying close contact with customers, patients, children or students contribute to the spread of COVID-19 and its severity is currently unknown.

Only a few studies have been published on the occupational risk of COVID-19, mainly focusing on disease severity or mortality. The first reports of occupational risk of COVID-19 are from Singapore in early February 2020 and showed that 25 locally transmitted cases were employed in tourism and trading [5]. Later British studies reported that essential workers such as personal service occupations and plant and machine operatives had a higher risk of severe COVID-19 than non-essential workers, which are believed to work more from home-office [6]. Also, in England, Wales and Sweden, occupations in sales and retail, transport (Swedish bus/taxi drivers) and catering (chefs) had raised mortality rates of COVID-19, whereas teachers had lower mortality rates [7,8]. Also, a number of outbreaks among meat workers and workers at abattoirs have been reported, underlining the potential of outbreaks in specific work settings [9].

An overview of the pattern of COVID-19, testing behavior, percent of the tested who were positive, and accompanying utilization of health care services in persons employed in a wide range of occupations is currently lacking. Improved knowledge of occupational testing behavior and risk would greatly contribute to informing authorities on testing regimens and whether certain activities in these sectors should be restricted in attempts to limit the spread of the virus. Most European countries including Norway are experiencing two waves of infection [10], one during spring 2020, and one during fall 2020, which due to the novelty of the virus and restrictions undertaken may be associated with different occupational risks. Thus, for the two periods of infection in Norway and including the entire Norwegian population aged 20-70 years, we aimed to study the occupational risk of confirmed COVID-19, associated testing behavior and hospitalizations for persons employed in health professions, education and teaching, trade, catering, travel, tourism and recreation industries.

## Methods

We utilized individual-level data from the BEREDT C19 register, which is a newly developed emergency preparedness register aiming to provide rapid knowledge of the spread of the SARS-CoV-2 virus and how spread, as well as measures to limit spread, affect the population’s health, use of health care services and health-related behaviors [11]. The register consists of electronic patient records from all hospitals in Norway (NPR), data from the Norwegian Surveillance System for Communicable Diseases (MSIS) including all laboratory tests, The Norwegian Population Registry and the Employer- and Employee-register, which are merged on the unique personal identification number that is provided every Norwegian resident at birth or upon immigration. Thus, BEREDT C19 and our study include the entire Norwegian population including immigrants. Data are updated daily (except for the Employer-Employee-register, which was updated on August 25^th^ 2020) and spans the whole of 2020. BEREDT C19 includes results for all positive and negative polymerase chain reaction (PCR) tests for SARS-CoV-2 as well as rapid antigen tests of every resident in Norway with dates of testing and test result, legally required to be reported from all laboratories to MSIS. The registration of positive cases is believed to be complete, while there may be lack of registration of negative tests before April 1^st^ 2020. BEREDT C19 also includes date of any hospitalization, with complete diagnostic codes. Occupation is reported in the Employer-Employee-register using Standard Classification of Occupations [12] for all residents in Norway. Thus, in the current study, our population included all Norwegian residents in their working age, here defined as age between 20 and 70 years on January 1^st^ 2020. Non-residents (like tourists, temporary workers and asylum applicants) were excluded. Institutional board review was conducted, and the Ethics Committee of South-East Norway confirmed (June 4th 2020, #153204) that external ethical board review was not required.

### Occupation

Occupation was registered with a 7-digit code in the Employer- and Employee-register according to the Standard Classification of Occupation (STYRK-98) [12]. To allow for international comparisons, we used a convert table to make the classification align with the Standard Classification of Occupations (ISCO-08 using 4-digit codes, i.e. corresponding to the Norwegian STYRK-08) [12, 13]. We selected common occupations with number of employees ≥1000 and number of contracted weekly work hours ≥1 for a reference week at the beginning of the pandemic (week 10). The occupations investigated in this study, classified as described in Table 1, usually imply direct contact with other people. Persons not registered with any of the STYRK-codes in Table 1 were classified as *everyone else in their working age (20-70 years)* and included persons with other occupations (i.e. unspecified occupation with an assumable low degree of contact with customers, patients, children or students). This category also included persons in the population register who had missing value on employment code for unknown reason (nonemployees like persons on disability pensions, work seekers, freelancers, self-employed and students).

**Table 1.** Occupations having direct contact with children, students, patients or customers.

| **Health occupations** |  |
| --- | --- |
|  | Code* |
| Physicians | 2211/<br>2212 |
| Nurses | 2221/<br>2223 |
| Dentists | 2261 |
| Physiotherapists | 2264 |
| **Teaching occupations** |  |
| Primary school teacher | 2341 |
| Early childhood educators/pre-school teachers | 2342 |
| Child/school care workers | 5311 |
| Secondary education teachers | 2330 |
| University & higher education teachers | 2310 |
| **Trade occupations** |  |
| Shop sales assistant | 5223 |
| Cleaners | 9112 |
| **Catering occupations** |  |
| Waiters | 5131 |
| Bartenders | 5132 |
| Food service counter attendant | 5246 |
| **Tourism & travel occupations** |  |
| Hotel receptionists | 4224 |
| Travel guides | 5113 |
| Travel attendants and travel stewards | 5111 |
| Transport conductors | 5112 |
| Bus and tram drivers | 8331 |
| Car, taxi and van drivers | 8322 |
| **Recreation & beauty occupations** |  |
| Fitness and recreation instructors and programme leaders | 3424 |
| Hair dressers | 5141 |

\*According to the International Standard for Classification of Occupation (ISCO-08 / STYRK-08)

### Outcomes

We studied two outcomes: 1) COVID-19, which was defined as either having a confirmed positive polymerase chain reaction (PCR) test for COVID-19, and/or by having ICD-10 diagnostic code U07.1 of confirmed COVID-19, and, 2) Hospitalization with confirmed COVID-19 [14]. Test criteria for COVID-19 initially included having severe disease, being in a risk group or being health personnel, later changing to include everyone with symptoms or having been in contact with persons with confirmed COVID-19 from the summer of 2020. Thus, we also studied testing behavior and split the analysis in two periods, before and after July 18^th^ 2020. At this date, the number of newly infected daily cases in Norway had decreased to about zero, and had been stable and low for several weeks in July before slowly rising again in the beginning of August [2-4]. We will refer to the two periods as the 1^st^ wave (including February 26^th^ – July 17^th^ 2020) and the 2^nd^ wave (including July 18^th^ – December 18th 2020).

### Statistical analyses

First, and for each of the occupation groups, we estimated the total number of confirmed COVID-19 cases per 1000 employees for the two waves of infection, as well as the percent of employees in an occupation that was tested for COVID-19 at least once, and the percent of the tested in each occupation that was positive (these descriptives may inform on an occupation group’s testing behavior and degree of underdiagnosing, i.e. to what extent the source of transmission is unknown). Second, we assessed the crude association between each of the exposure occupation group (i.e. a categorical variable including the 22 categories, one for each occupation) and the outcome confirmed COVID-19 (yes/no) using logistic regression separately for each of the waves and reporting odds ratios (OR). Third, we assumed that several potential covariates may confound the association between occupation and wave-specific COVID-19 outcome, and we adjusted for the following covariates in three multivariate logistic regression models: 1) age and sex, 2) age, sex and number of negative PCR-tests, and 3) age, sex, number of negative PCR-tests, country of birth, and mother’s country of birth. Given the large number of observations, we implemented the covariates as categorical variables (5 age categories: 20-29, 30-39, 40-49, 50-59, 60-70 years; 5 testing categories: 0, 1, 2, 3 and 4 or more negative tests; 7 categories for own and maternal countries of birth (in separate variables): Born in Norway, rest of Europe, Asia, Africa, Latin America, North America or Oceania, or unknown.

We set everyone else in their working age (20-70 years) to be the reference category in all analyses. We repeated both the descriptive and logistic regression analyses for each of the eleven administrative counties of Norway. Finally, we repeated the analyses using hospitalization with COVID-19 as outcome, however, due to a low number of hospitalizations for several occupation groups, we did not separate these analyses on the 1^st^ and 2^nd^ wave, nor did we study these outcomes per county. The statistical software used was STATA MP v.16.

## Results

We studied all 3 559 694 persons aged 20-70 years living in Norway on January 1^st^ 2020 with mean (SD) age 44.1 (14.3) years, consisting of 51% men. Of these, 74.2% had birth country Norway (50% of those not born in Norway, were born in another European country) and 24.4% were nonemployed or not registered with any occupation. By December 18^th^ 2020, a total of 30 003 (0.8%) had contracted COVID-19, of which 1 550 (5.2%) had been hospitalized with COVID-19. The percent tested by occupation, as well as the percent of the tested who were positive by occupation, are reported in Figures 1 and 2, respectively. Generally, a high percent of health personnel and teachers were tested, while the percent of the tested who were positive was high among employees in catering, travel and tourism occupations (Figure 1 and 2). There were only minor regional differences between the occupations in testing/testing positive patterns (Supplementary (S-)Figure A-K). The proportions with COVID-19 and related hospitalization per occupation are reported in Table 2.

**Table 2.** Proportion per occupation with confirmed COVID-19 per 1000 employees, and proportion per occupation hospitalized with COVID-19 per 1000 infected employees.

|  | COVID-19<br>1st wave<br>(Feb 26th-July 17th<br>2020) | COVID-19<br>2nd wave<br>(July 18th-Oct 20th<br>2020) | Hospitalized<br>with<br>COVID-19<br>Total period<br>(Feb 26th -<br>Dec 18th<br>2020) |
| --- | --- | --- | --- |
| Everyone in their working age<br>(20-70 yrs) | 2 | 7 | 48 |
| Health occupations |  |  |  |
| Physicians | 7 | 7 | 24 |
| Nurses | 6 | 8 | 38 |
| Dentists | 6 | 5 | 244 |
| Physiotherapists | 3 | 4 | 14 |
| Teaching occupations |  |  |  |
| Primary school teacher | 1 | 8 | 28 |
| Early childhood educators | 1 | 7 | 37 |
| Child care workers | 2 | 10 | 27 |
| Secondary education teachers | 1 | 6 | 40 |
| University & higher education<br>teachers | 2 | 6 | 43 |
| Trade occupations |  |  |  |
| Shop sales assistant | 2 | 9 | 23 |
| Cleaners | 2 | 10 | 47 |
| Catering occupations |  |  |  |
| Waiters | 2 | 16 | 26 |
| Bartenders | 2 | 17 | 31 |
| Food service counter attendant | 3 | 17 | 35 |
| Tourism & travel occupations |  |  |  |
| Hotel receptionists | 1 | 7 | 26 |
| Travel guides | 1 | 8 | 0 |
| Travel attendants and travel<br>stewards | 2 | 11 | 0 |
| Transport conductors | 1 | 11 | 87 |
| Bus and tram drivers | 6 | 11 | 95 |
| Car, taxi and van drivers | 4 | 13 | 77 |
| Recreation & beauty occupations |  |  |  |
| Fitness and recreation instructors<br>and programme leaders | 1 | 7 | 0 |
| Hair dressers | 1 | 8 | 31 |

\*According to the International Standard for Classification of Occupation (ISCO-08 / STYRK-08)

**Figure 1.**
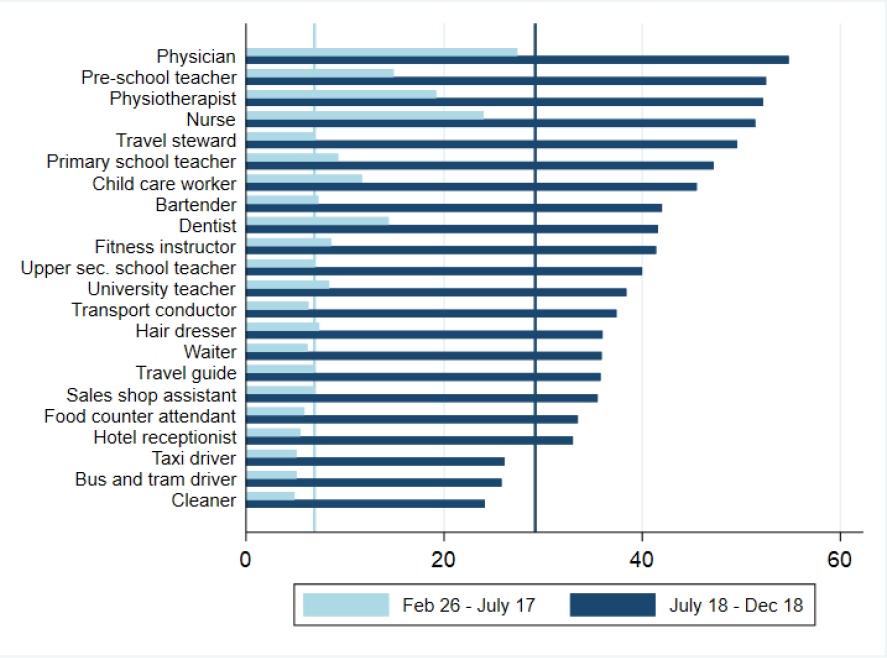
Percent of employees who were tested at least once in the given period, by occupation. Vertical line indicates the mean in the reference group (everyone else aged 20-70).

**Figure 2.**
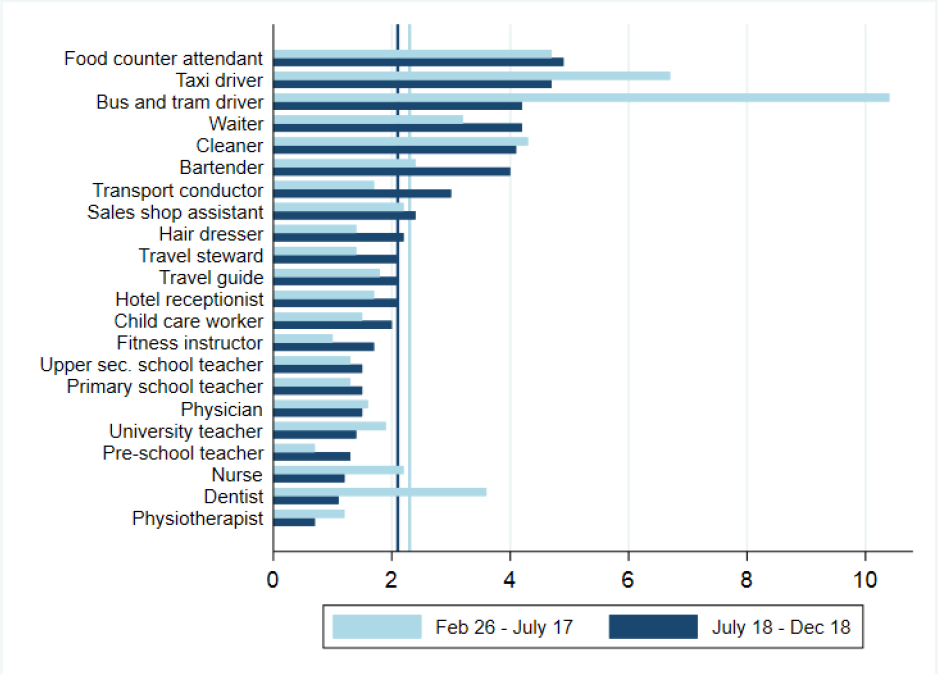
Percent of the tested employees for whom at least one test was positive in the given period, by occupation. Vertical line represents the mean in the reference group (everyone else aged 20-70).

### Risk of confirmed COVID-19, 1^st^ wave (Feb 26^th^- July 17^th^ 2020)

Persons employed as dentists, nurses, physicians, physiotherapists, bus and tram and taxi drivers had ∼1.1-4 times the odds of confirmed COVID-19 during the first wave of infection when compared to everyone else aged 20-70 and jointly adjusting for age, sex, testing behavior and continent of birth (Figure 1). In contrast, teachers of children and students at any age, child and school care workers, as well as bartenders, waiters, sales shop assistants, cleaners, fitness instructors, hair dressers, hotel receptionists, travel guides and transport conductors had no increased risk, or even a reduced risk of COVID-19 when compared to everyone else aged 20-70 (Figure 3). Generally, point estimates were closer to 1 in adjusted models when compared to the crude model without covariates (Figure 3). A large deviance between the estimates from the different regression models for an occupation may imply that other factors than occupation (i.e. age, sex, testing regimen/tendency and/or immigrant status) explains in whom COVID-19 is detected. In analyses stratified by region, there was considerable uncertainty, however health personnel tended to have a higher OR than everyone else aged 20-70 (S-figure A-K).

**Figure 3.**
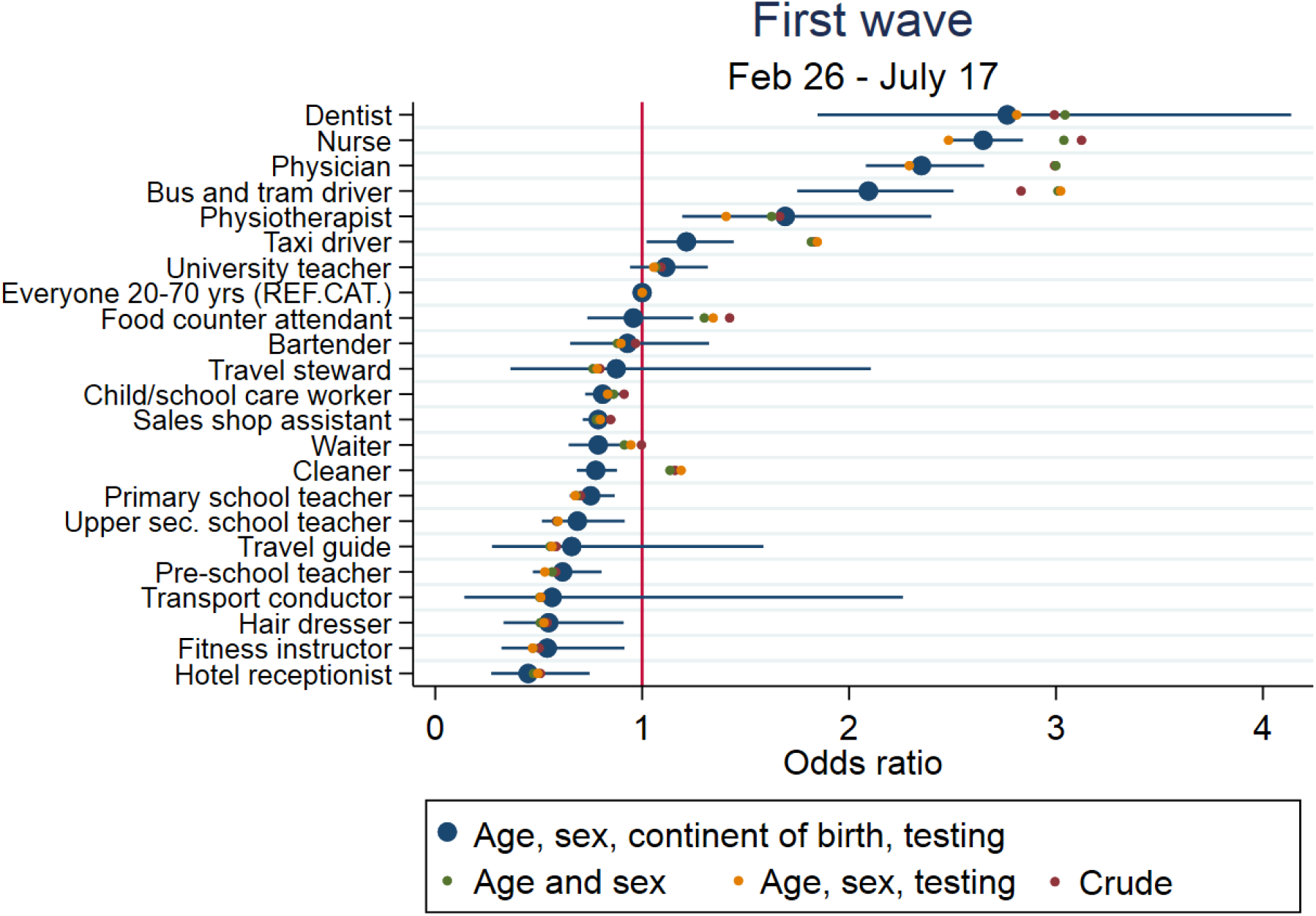
The odds (95% confidence interval) of COVID-19 during the 1^st^ wave of infection in Norway (Feb 26^th^-July 17^th^ 2020), with different levels of adjustment. Everyone else aged 20-70 was the reference category (OR==1, vertical red line).

### Risk of confirmed COVID-19, 2^nd^ wave (July 18^th^- December 18^th^ 2020)

The pattern of occupational risk of COVID-19 was different for the 2^nd^ wave of infection than for the 1^st^ wave of infection. In the 2^nd^ wave, transport conductors, bartenders and travel stewards had ∼1.1-3 times the odds of COVID-19 when compared to everyone else aged 20-70 and jointly adjusting for age, sex, testing behavior and continent of birth (Figure 4). A range of occupations had moderately increased odds (OR∼1.1 to OR∼1.5): waiters, food service counter attendants, bus, tram- and taxi drivers, child/school care workers, primary-, pre-school and upper secondary teachers and hair dressers, physicians and sales shop assistants when compared to everyone else aged 20-70 (Figure 4). Also, other health personnel had a lower to only moderately increased odds in the second wave when compared to everyone else aged 20-70 (Figure 4). Again, point estimates were often closer to 1 in adjusted analyses when compared to crude analyses (Figure 4). As an example, the crude OR of ∼1.5 for cleaners was reduced to 1 in analyses jointly adjusted for age, sex, testing activity and continent of birth (Figure 1, Figure 4).

**Figure 4.**
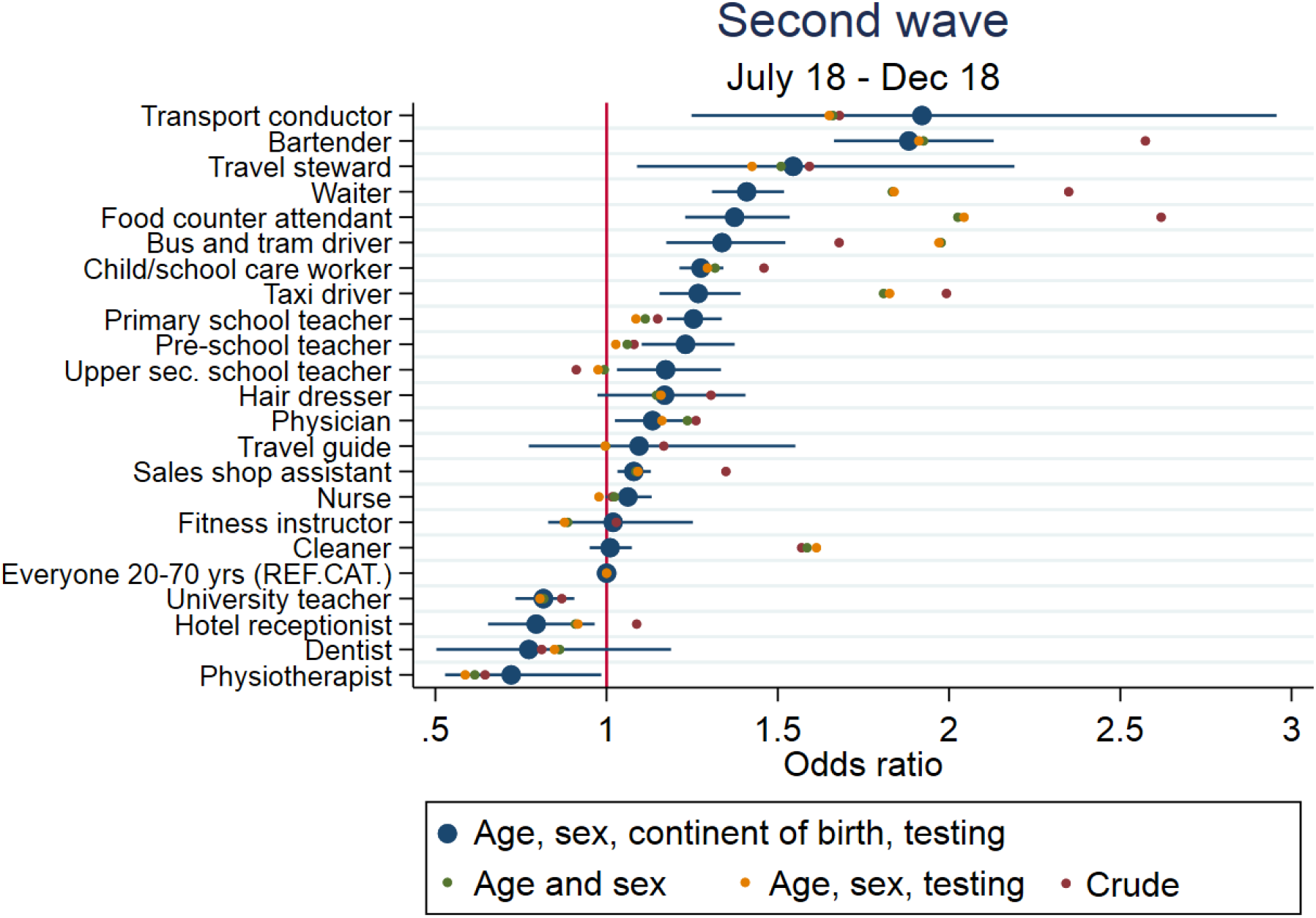
The odds (95% confidence interval) of COVID-19 during the 2^nd^ wave of infection in Norway (July 18^th^ – December 18^th^ 2020), with different levels of adjustment. Everyone else aged 20-70 was the reference category (OR==1, vertical red line).

There were large variations in estimates and considerable uncertainty in the occupational risks by region. Most importantly, teachers had a high test activity in close to all counties (S-Figure A-K), and their estimated occupational risk tended to be up to doubled (OR ∼2) only in a few counties, i.e. in counties Oslo (child/school care worker, pre- and primary school teacher) and Innlandet (pre-school teacher) (S-figure A-K).

### Risk of hospitalization with COVID-19

None of the included occupations had any particularly increased risk of severe COVID-19, indicated by hospitalization, when compared with everyone else infected aged 20-70 (Figure 5), apart from dentists, who had a ∼7 (2-12) times increased odds ratio. However, for several occupations, no hospitalizations were observed, confidence intervals were wide and all analyses should be interpreted with caution due to the small number of COVID-19 hospitalizations.

**Figure 5.**
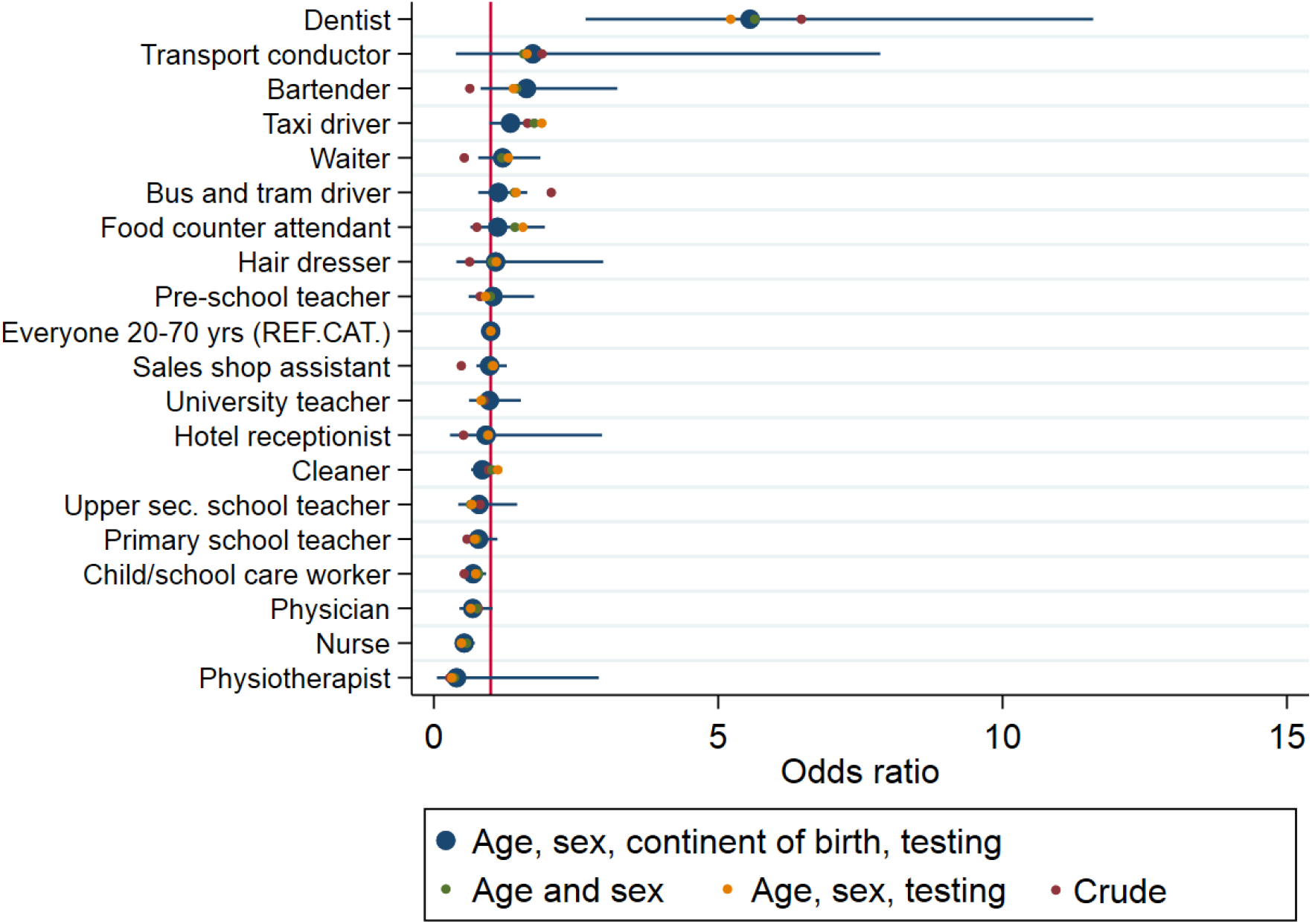
The odds (95% confidence interval) of being hospitalized with COVID-19 during the 1^st^ and 2^nd^ wave of infection in Norway (Feb 26^th^-December 18^th^ 2020) with different levels of adjustment. Everyone else aged 20-70 with COVID-19 was the reference category (OR==1, vertical red line).

## Discussion

Studying the entire Norwegian population, we report a different pattern of occupational risk of COVID-19 for the 1^st^ and the 2^nd^ wave of infection. Importantly, nurses, physicians, dentists, physiotherapists, bus and tram and taxi drivers had the highest risk of confirmed COVID-19 during the 1^st^ wave of infection, which shifted to bartenders, transport conductors and travel stewards during the 2^nd^ wave of infection (compared to everyone else in their working age 20-70). Teachers had a moderately increased risk of COVID-19 during the 2^nd^ wave of infection, however, the risk was up to double in counties Oslo and Innlandet. We found indications that occupation may be of limited relevance for the risk of severe COVID-19, here studied as hospitalization with the disease.

We believe this report is the first to show the COVID-19 risks of specific occupations for the entire working population and for everyone diagnosed. Existing reports have considered the associations in smaller populations, have used broad categories of occupations and/or have considered only severe, hospital-confirmed COVID-19 or mortality [5-8]. Here, we study everyone with negative and positive polymerase chain reaction (PCR) test for SARS-CoV-2 in Norway in addition to hospital-confirmed COVID-19 as well as hospitalizations with COVID-19. For studying occupations, we use the internationally well-known ISCO-codes with four digits, and apply logistic regression models, making analyses reproducible and comparable when repeated in other countries or in other study samples.

Considering that workers both may become infected through their occupation but may also spread the virus to their customers, patients or pupils/students, our findings may have implications for pandemic policy. First, our findings give reason to believe that bartenders, waiters, travel stewards, bus, tram and taxi drivers had a higher risk of infection than other occupation groups in the 1^st^ and/or 2^nd^ wave, and they also typically have contact with many different people in their work possibly exposing many people if they are not aware that they are infected. Additionally, these occupation groups had a low frequency of testing and a high percentage positive tests among the tested. These findings may be of relevance for the future considerations of testing regimens by occupations, for the implementation of restrictions and/or for the use of face masks in certain occupational settings.

We also found indications that child/school care workers, teachers in pre-school, primary school and upper secondary school may be at a moderately increased risk of COVID-19 in the second period (OR ∼1.25). In analyses stratified by region, teachers or child/school care workers had an even higher odds in two counties (Innlandet, Oslo), with OR ∼2. This pattern may partly be explained by a high testing tendency in these occupation groups and low percentage of positive among the tested (Figure 1, Figure 2 and S-Figures). However, considering the increasing transmission particularly during fall 2020 [4], we cannot exclude that teachers have been infected at work, by their pupils and/or by their colleagues. A recent Norwegian study found low risk of transmission from school children to adults during the same period (fall 2020) [15], which may imply that teachers are infected by each other in their work settings, rather than by their pupils. The OR for teachers were generally lower in the first period, which may be explained by schools mainly being closed due to restrictions or summer holidays.

Except for our analyses of hospitalization, we chose to divide our analyses in two periods, the 1^st^ and 2^nd^ waves [10]. An important potential explanation for the differing findings in the 1^st^ and 2^nd^ wave may be differences in test criteria in Norway through the year, which changed from including only those with severe disease, at risk, and/or health personnel before summer to include everyone with mild symptoms and several other test indications after summer. These differences in test criteria may also explain why health personnel were at increased risk during the 1^st^ wave but not the 2^nd^ wave. Indeed, we show that health personnel have had a high test frequency throughout the pandemic (Figure 1, 2). However, it is also possible that health personnel have implemented better infection control measures, resulting in fewer nurses, dentists etc. being infected as the pandemic progressed. Along this line, our work raises important hypotheses: Although we had few cases and considerable uncertainty in our analyses of hospitalization with COVID-19, the considerably increased risk of severe COVID-19 for dentists, calls for further research of the relevance of viral load or infectious dose in causing severe COVID-19. Future research should also detail the association between testing regimens, infection risk and disease severity across types of health/medical occupations.

Another issue of importance to the interpretation of our findings is that 24% of the working age population could not be categorized using available registry data, i.e. they may be everything from students and freelancers to those unemployed and disability pensioned. As an example, the persons infected during the 2^nd^ wave of infection were younger and likely consisted of more students when compared to persons infected in the 1^st^ wave of infection [2-4]. This may be due to the fact that the younger part of the population typically have less severe symptoms, and therefore probably tended not to be tested during the first wave of the epidemic. The students, typically aged 20-25, may more often than those aged ≥30, have no occupation, and/or more often have part-time work as bartenders, waiters, food counter attendants, child care workers, sales shop assistants etc., potentially explaining our results. The non-employed might also be on disability pensions, typically due to poor health and potentially at greater risk of severe COVID-19, potentially explaining why our findings indicate limited occupational risk of hospitalization with COVID-19. In total 12% of non-elderly adults in Norway are on disability pensions. Also, the proportions fully or partially retired increases from ∼ 0% to ∼ 95% between age 60 and age 70 [16], and they may be exposed to a steeply declining occupational risk.

Some important limitations should be mentioned. First, we cannot exclude that other factors than the occupation in question explain infection and hospitalization risks in our study. As an example, persons in full-employment may be at greater risk of COVID-19 than persons in part-time employment. Also, we cannot be sure we have sufficiently adjusted for other risk factors related to e.g. country of birth, residential area, risky behavior and health literacy, which may be of particular relevance to our analyses of hospitalization [8]. Further, it is possible that employees working and living close together in small areas (more typical for big cities) may be infected by each other rather than by the customers/children/patients they meet [17]. Indeed, point estimates and their 95% CI were generally lowered in adjusted compared to crude analyses, suggesting that occupation and our outcomes are partly explained by sociodemographic factors. Our stratified analyses may shed further light on the differences in occupational risk in rural and urban areas (S-Figure A-K). However, we had sparse data in several of the counties studied, and the county-specific analyses should be interpreted with caution due to low numbers. Another potential limitation is the validity of negative tests in the beginning of the pandemic, before April 1^st^ 2020. Finally, we converted the Norwegian occupation classification to ISCO-08 and some of the occupations (0.3%) were lost as they did not convert to the international system [12, 13]. The reference category was calculated using STYRK-98.

In conclusion, we show that nurses, physicians, dentists, physiotherapists, bus, tram and taxi drivers had the highest risk of confirmed COVID-19 during the 1^st^ wave of infection, which shifted to bartenders, waiters, travel stewards and transport conductors during the 2^nd^ wave of infection. Teachers had a moderately increased risk of COVID-19. Our findings may be of relevance to increase the understanding of risk and transmission settings for COVID-19 in order to contribute to more targeted measures to decrease transmission of COVID-19 in public settings.

## Data Availability

Data are not publicly available.

## Acknowledgements

We would like to thank the Norwegian Directorate of Health, in particular Director for Health Registries Olav Isak Sjøflot and his department, for excellent cooperation in establishing the emergency preparedness register. We would also like to thank Gutorm Høgåsen, Ragnhild Tønnessen and Anja Elsrud Schou Lindman for their invaluable efforts in the work on the register. The interpretation and reporting of the data are the sole responsibility of the authors, and no endorsement by the register is intended or should be inferred. We would also like to thank everyone at the Norwegian Institute of Public Health who has been part of the outbreak investigation and response team. In particular, our acknowledgements to Margrethe Greve-Isdahl and Pål Suren for support in interpretation and communication of results.

## Author contribution

Karin Magnusson had access to all of the data in the study and takes full responsibility for the integrity of the data and the accuracy of the data analysis. Karin Magnusson drafted the manuscript. Karin Magnusson and Fredrik Methi performed statistical analyses. Fredrik Methi, Karin Nygård, Line Vold and Kjetil Telle contributed with acquisition of data, conceptual design, analyses and interpretation of results. All authors contributed in drafting the article or critically revising it for important intellectual content. All authors gave final approval for the version to be submitted.

