## Supplementary material for "Occupational risk of COVID-19 in the 1^st^ vs 2^nd^ wave of infection": S-Figure

### Supplementary file for the paper

### Occupational risk of COVID-19 in the 1st vs 2nd wave of infection

By Magnusson et al., 2020

Supplementary Figure A: Agder p. 2

Supplementary Figures B-C: Innlandet and Møre og Rmsdal p. 3

Supplementary Figures D-E: Nordland and Oslo p. 4

Supplementary Figures F-G: Rogaland and Troms og Finnmark p. 5

Supplementarly Figures H-I: Trøndeland and Vestfold og Telemark p. 6

Supplementary Figures J-K: Vestland and Viken p. 7

Supplementary Figure A-K. Percentage tested, percentage of the tested who were positive and logistic regression analyses of confirmed SARS-CoV-2 by occupation and county, for the 1st and 2nd wave of infection. Logistic regression models adjustment with following covariates (*crude* includes no covariates):

- Age, sex, continent of birth, mother's continent of birth and testing
- Age and sex
- Age, sex and testing
- Crude

A. Agder

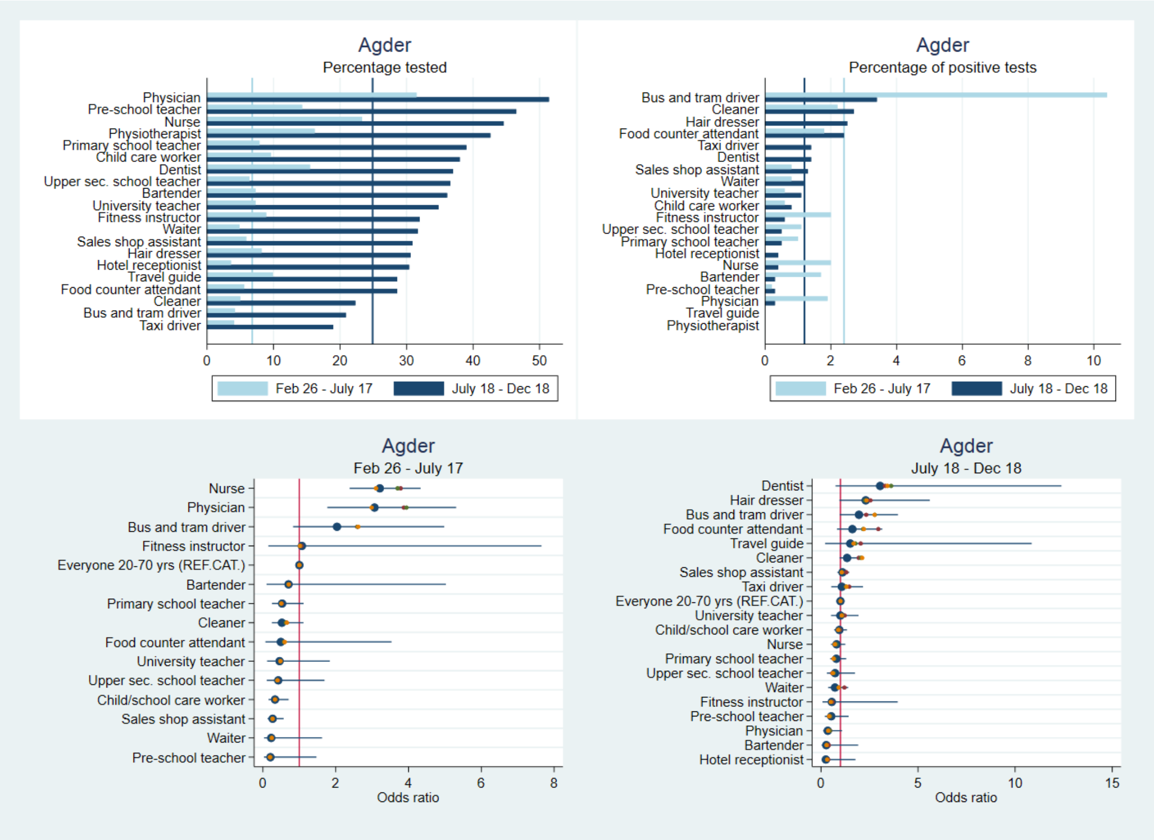

#### B. Innlandet

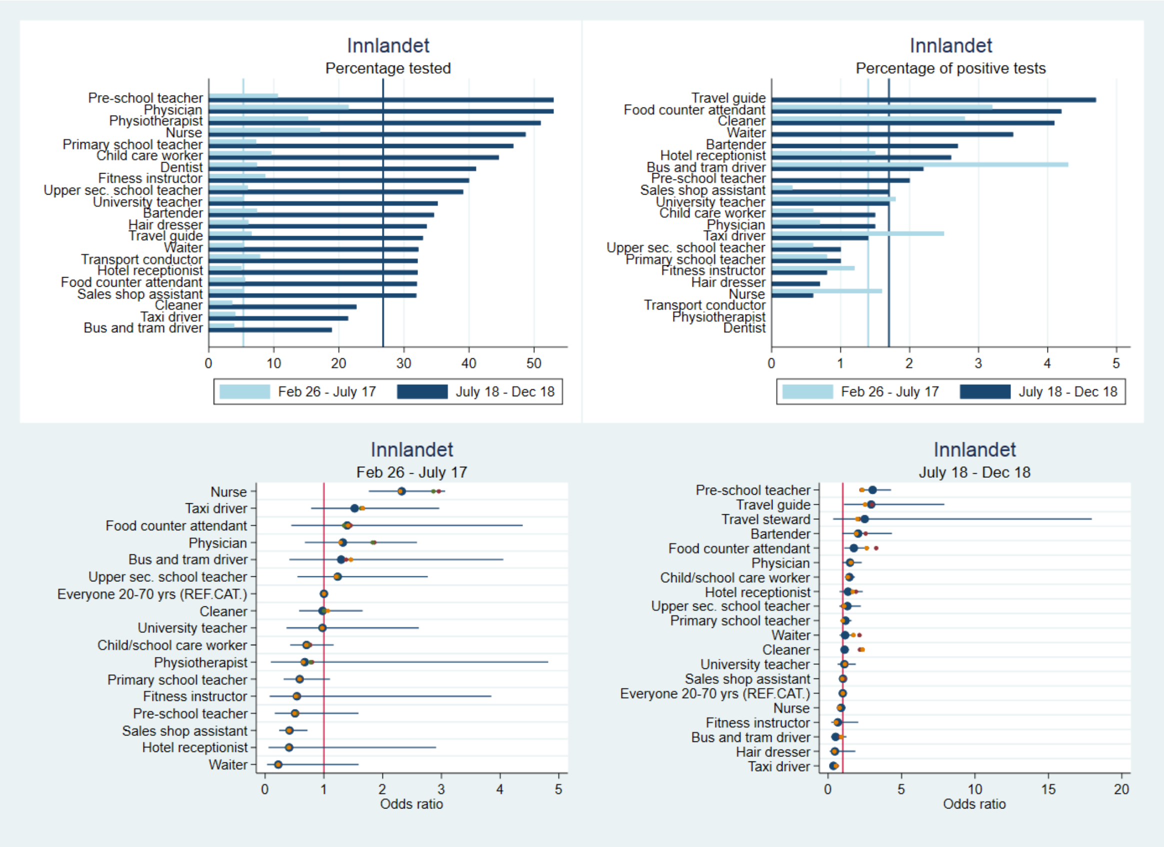


#### C. Møre og Romsdal

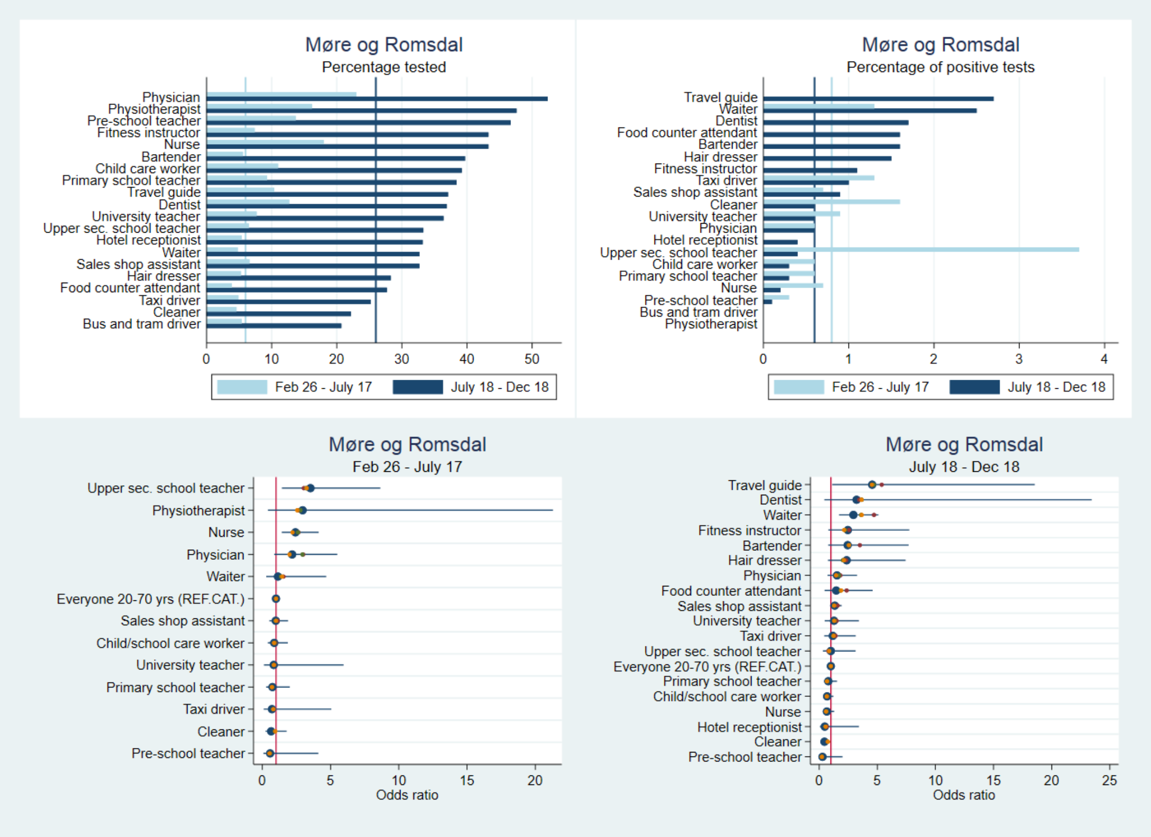

D. Nordland

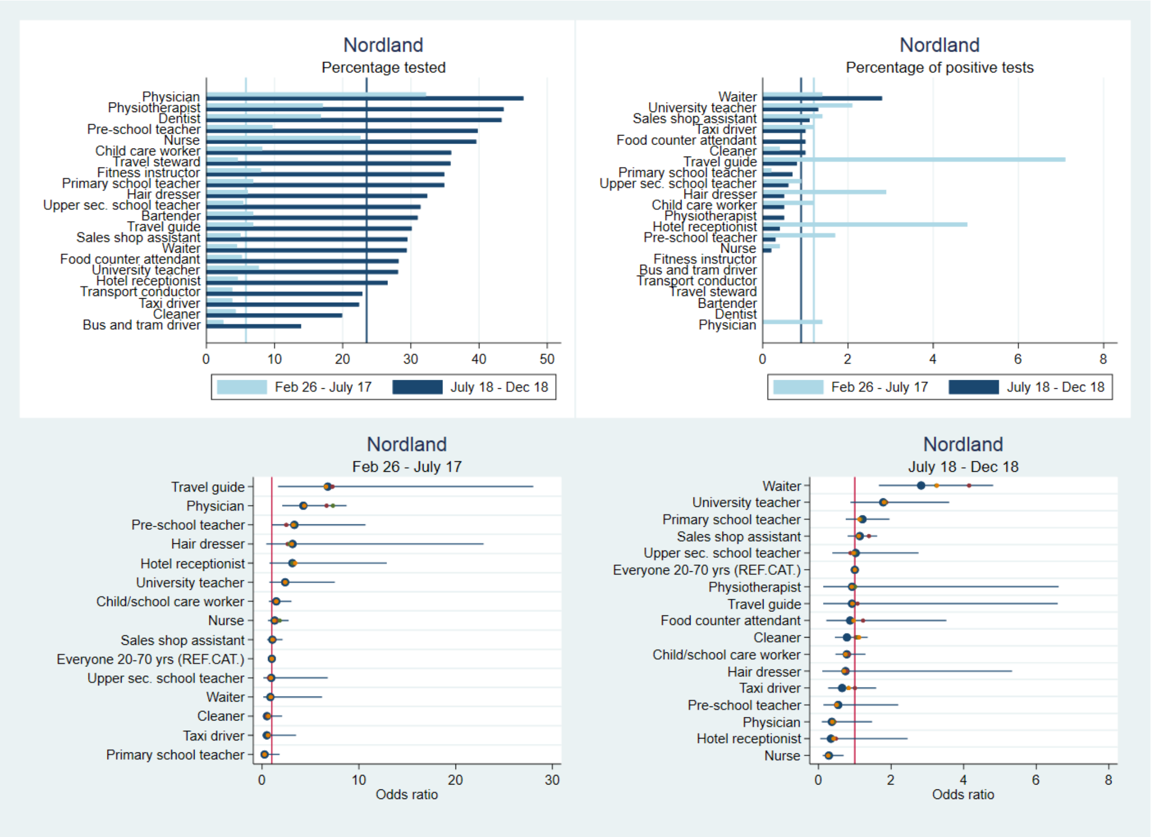


E. Oslo

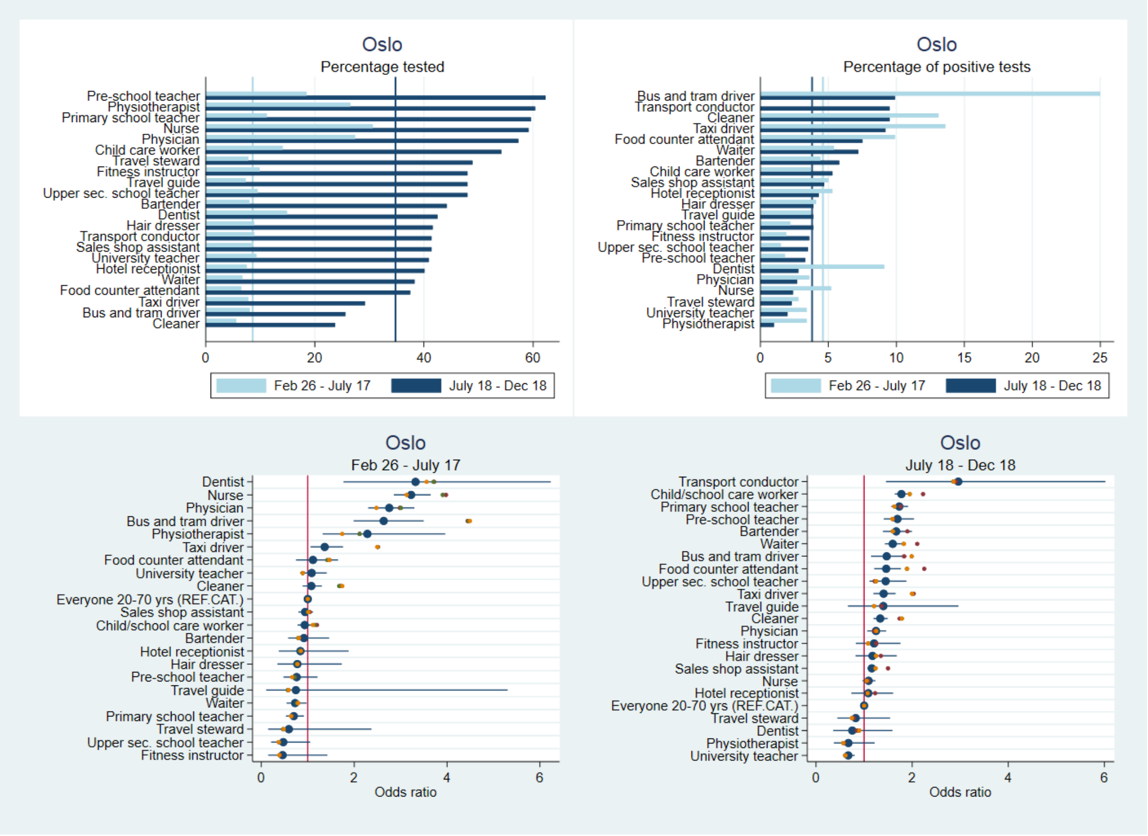

#### F. Rogaland

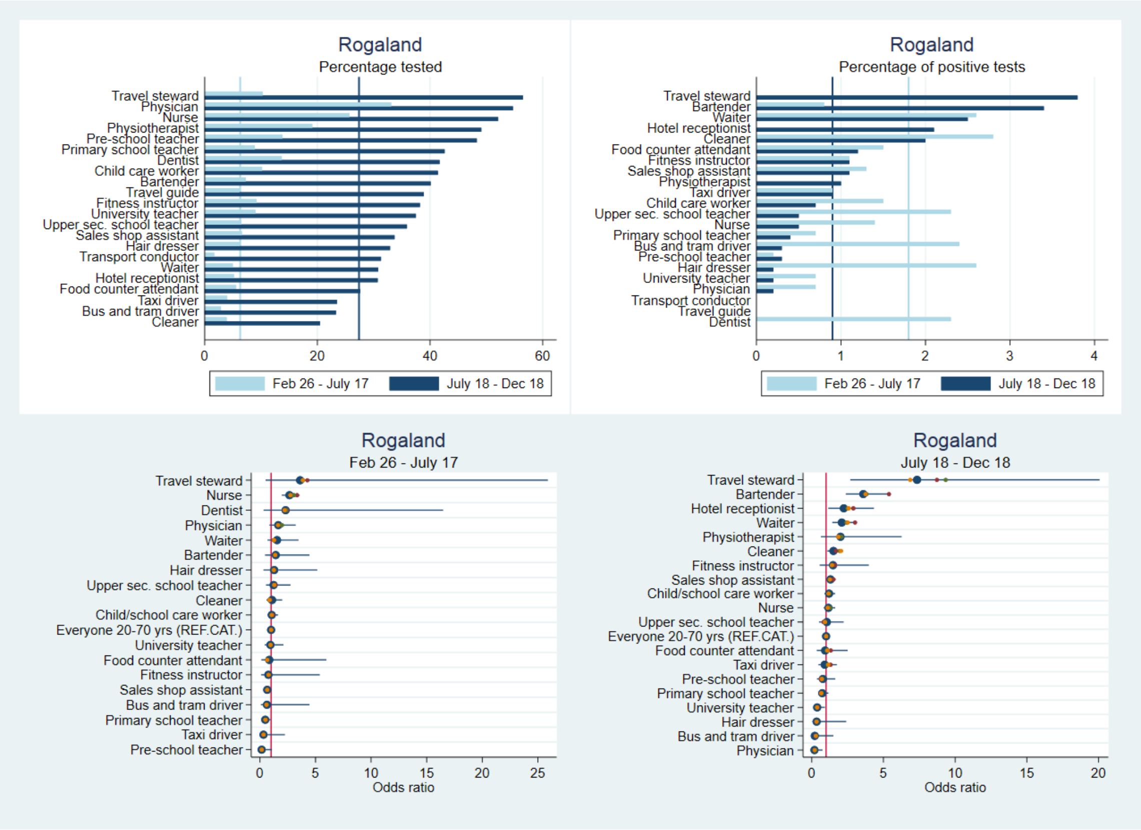


#### G. Troms og Finnmark

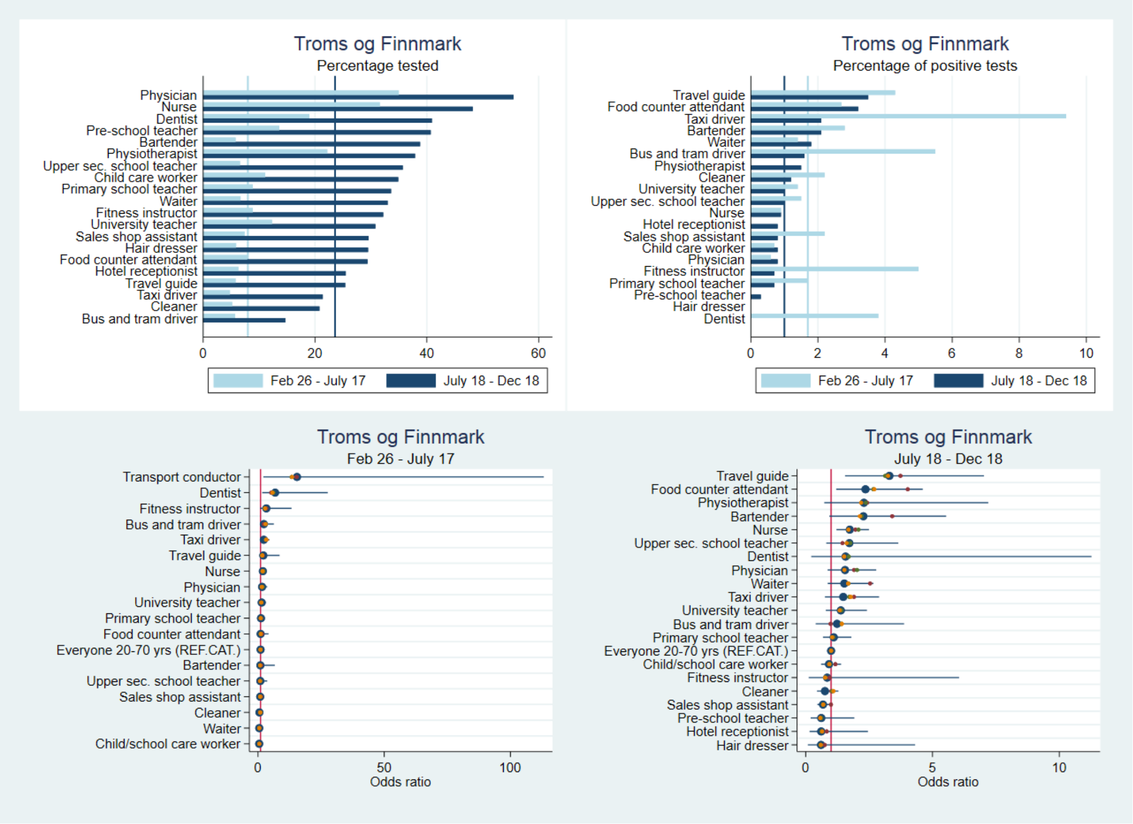

#### H. Trøndelag

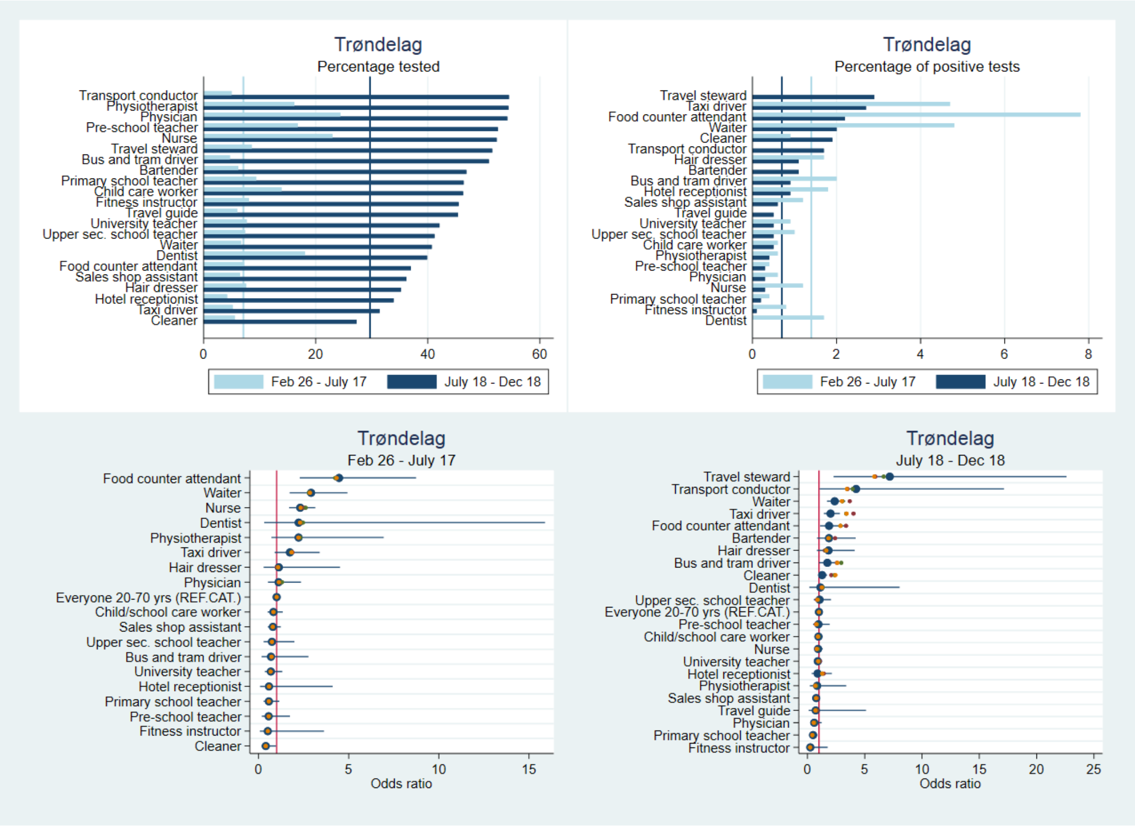


#### I.Vestfold og Telemark

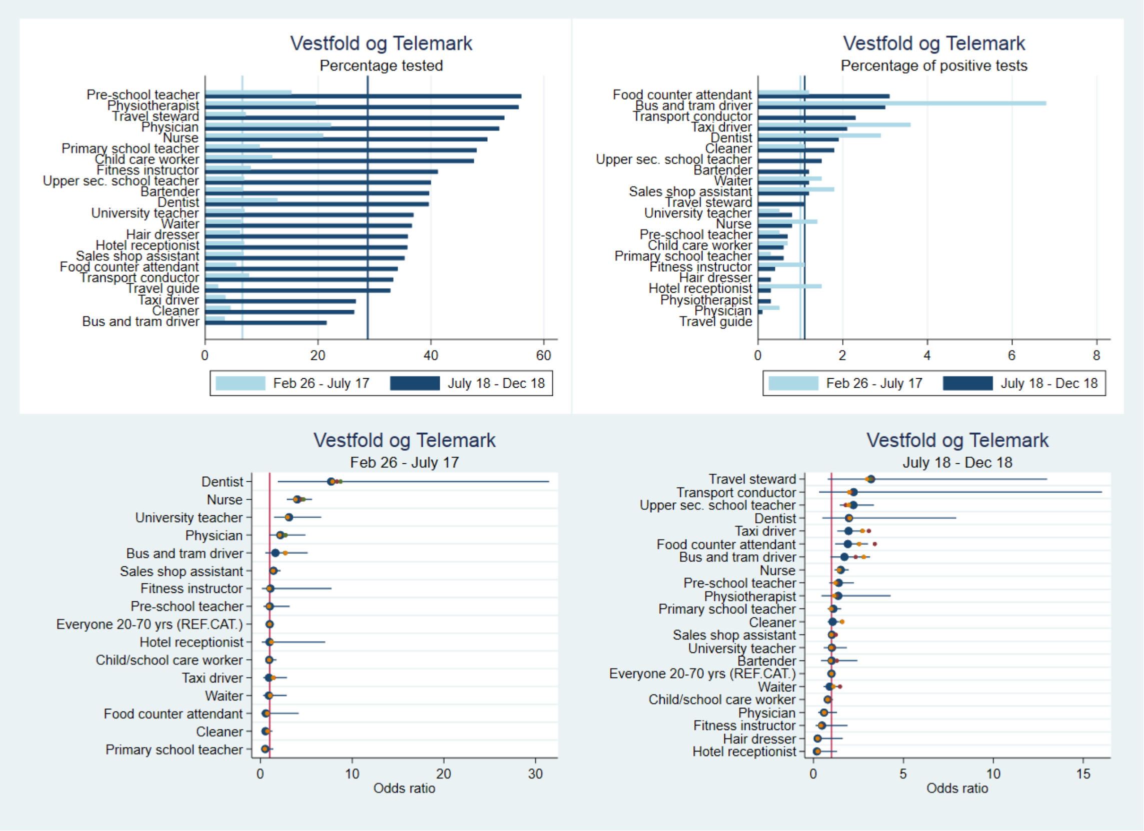

J. Vestland

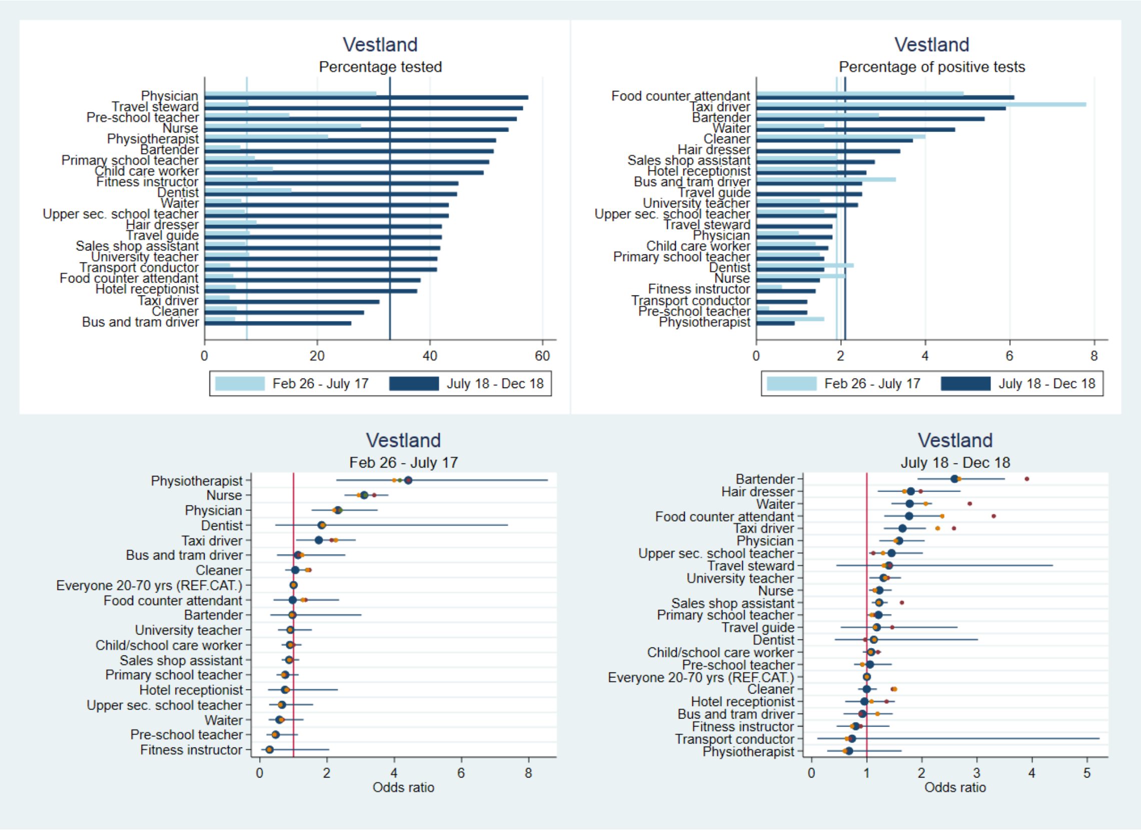

K. Viken

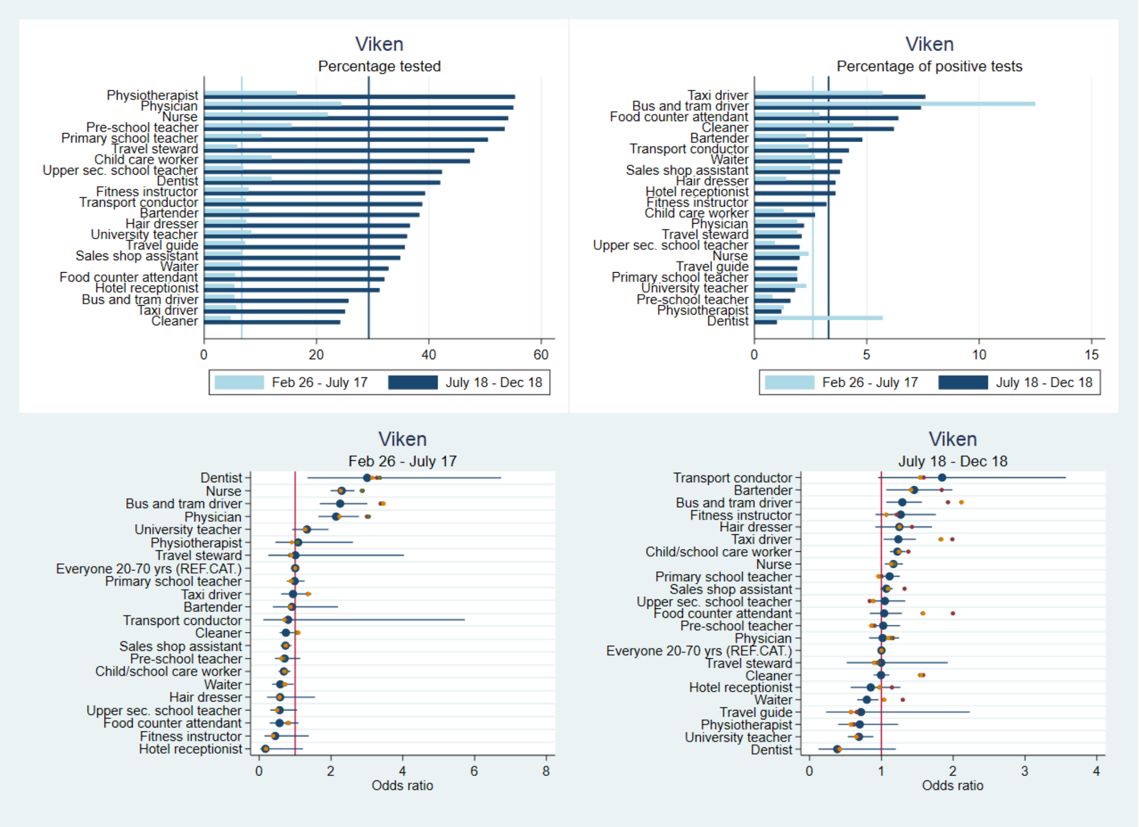
